# Factors Associated with Open Access Publishing Costs in Oncology Journals

**DOI:** 10.1101/2022.05.10.22274921

**Authors:** Alex Koong, Ulysses G Gardner, Jason Burton, Caleb Stewart, Petria S Thompson, Clifton D Fuller, Ethan B Ludmir, Michael K Rooney

**Affiliations:** Department of Radiation Oncology, MD Anderson Cancer Center, Houston, TX; Kettering Health, Dayton, OH; Dartmouth-Hitchcock Medical Center, Lebanon, NH; Unity Health, Searcy, AR; St Mary’s Medical Center, San Francisco, CA

**Keywords:** open access, article processing charges, academic publishing

## Abstract

**Background:** Open access (OA) publishing represents an exciting opportunity to facilitate dissemination of scientific information to global audiences. However, OA publication is often associated with significant article processing charges (APCs) for authors, which may thus serve as a barrier to publication.

**Methods:** We identified oncology journals using the SCImago Journal & Country Rank database. All journals with an OA publication option and APC data openly available were included. We searched journal websites and tabulated journal characteristics, including APC amount (USD), OA model (hybrid vs full), 2-year impact factor (IF), H-index, number of citable documents, modality/treatment specific (if applicable), and continent of origin. We generated a multiple regression model to identify journal characteristics independently associated with OA APC amount.

**Results:** Of 367 oncology journals screened, 251 met final inclusion criteria. The median APC was 2957 USD (IQR 1958-3450). On univariable testing, journals with greater number of citable documents (p<0.001), higher IF (p < 0.001), higher H-index (p < 0.001), and those using the hybrid OA model (p < 0.001) or originating in Europe/North America (p < 0.001) tended to have higher APCs. In our multivariable model, number of citable documents, IF, OA publishing model, and region persisted as significant predictors of processing charges.

**Conclusions:** OA publication costs are greater in oncology journals that publish more citable articles, utilize the hybrid OA model, have higher IF, and are based in North America or Europe. These findings may inform targeted action to help the oncology community fully appreciate the benefits of open science.

## Introduction

Open access (OA) publication grants any individual permission to view published scientific articles without cost, thereby allowing free public access of content regardless of geography or consumer affiliation.(1) This contrasts with traditional subscription-based publishing which either require institutional subscriptions to grant journal access or require fees for access to individual articles. Various OA publication models exist, although in general can be considered as two broad categories. “Hybrid OA” refers to subscription-based journals that allow authors an option of making individual articles OA whereas “full OA” journals publish exclusively OA content.(2)

OA publishing allows for advancements in science and medicine to be adopted at faster rates and promotes the democratization of knowledge. As such, there has been a rapid growth of OA publishing, with an estimated average growth of 30% in number of published articles per year since the early 1990s.(3) Clinicians and patients with complete access to relevant studies can make more informed health decisions, rather than attempting to base their choices solely on the subset of studies from which they have access. Many journals require authors to pay article processing charges (APCs) in order to publish an OA manuscript. These fees can be significant, in the ranges of thousands of US dollars (USD), and thus may inadvertently serve as a barrier to OA publication, particularly for authors with limited personal or institutional resources.

In this study, we sought to identify and characterize journal-level factors associated with APCs in oncology journals. We hypothesized that journals with higher impact factors (IF), those based in the United States, and those adopting the hybrid OA model would higher APCs. Our results could be used to inform policy level change aimed at promoting equity and equality in access to oncologic research.

## Methods

### Journal identification

The SCImago Journal & Country Rank (SJR) database (https://www.scimagojr.com) was queried on August 19, 2020 to identify oncologic journals. The following search parameters were used: subject area: “Medicine”; category: “Oncology”; region/country: “All regions/countries”; types: “Journals”; Year: “2019.” Resulting journals were screened according to OA publishing status. Journals with an OA publishing option (hybrid or full) and APC data available via their website were included. Journals that were discontinued or written in non-English language were excluded for this analysis. Journals with less than 10 published articles per year were also excluded. Institutional review board approval was not necessary for this study and was waived accordingly. Informed consent was not required as the included information is publicly available.

### Journal evaluation

For each journal meeting inclusion criteria, the journal’s website was manually searched, and the following data points were tabulated: APC (in USD), OA model (hybrid or full), two-year IF, H-index, number of citable documents, modality specificity (yes or no), treatment site-specificity (yes or no), and continent of origin (North America, Europe, or other). A journal was considered modality-specific if its stated mission and scope was to publish content related to a particular oncologic treatment modality (e.g. surgery, radiation therapy, etc). All APCs were converted to USD equivalents for final analyses. An audit of a random 10 percent sample of the data was independently performed by two authors (MKR and UGG) to ensure data accuracy, precision, and reproducibility.

### Statistical analysis

Data were summarized using descriptive statistics. Pearson correlation was used to identify associations between continuous variables. Wilcoxon rank sum and Kruskal-Wallis testing were utilized to identify differences in continuous variables. For factors with significant associations on univariable testing, a multiple regression model was created to identify factors independently associated with APCs. For collinear factors, only one was utilized in the multivariable model. Log transformation was used for right-skewed predictive variables. All analyses were performed with R 4.0.3 (R Foundation for Statistical Computing, Vienna, Austria).

## Results

Of 367 oncology journals identified in the SCImago database, 251 met final inclusion. The most common reasons for exclusion were journals not having an OA publishing option or not having detailed information regarding APCs on the journal website. The majority of journals (62%) adopted the hybrid OA publication model and were based in Europe (47%) or North America (35%). The median (interquartile range (IQR)) APC for all journals was 2957 (1958-3540) USD. Twenty-five (10%) journals had APCs greater than 4000 USD. There were 10 journals (4%) which offered OA publication with no publication charge. The median number of citable documents per year was 308; however, there was significant variation in publication volume across evaluated journals. Fourteen percent of journals published fewer than 100 articles per year while 11% published at least 1000 citable articles per year.

The Table summarizes journal baseline characteristics and the results of the univariable and multivariable models to identify predictors of publication costs. Increased number of citable documents (P=0.006), higher IF (P<0.001), use of the hybrid OA model (P<0.001), and North American origin (P<0.001), were independently associated with increased APCs according to the multivariable model. Beta coefficients are provided in the Table to estimate the effect of various independent variables on APCs. For every 10-fold increase in number of citable documents and journal IF, there is an estimated increase of 367 USD and 1144 USD in APC, respectively. The use of the hybrid OA model was associated with an increase of 991 USD in APCs. Furthermore, compared to European journals, North American journal APCs were predicted to be 838 USD higher.

**Table:** Univariable and multivariable models to identify factors associated with journal article processing charges.

|  | **Univariable Model** |  | **Multivariable Model** |  |
| --- | --- | --- | --- | --- |
| **Characteristic** | **Median (IQR) or Count (%)** | ***P*-value** | **Beta (SE) USD** | ***P*-value** |
| APC (USD) | 2957 (1958-3540) | NA |  |  |
| Mean Number of Citable Documents per Year<sup>a</sup> | 308 (140-536) | <0.001 | 367* (133) | 0.006 |
| 2 year Impact Factor<sup>a</sup> | 2.8 (2.0-4.1) | <0.001 | 1144* (177) | <0.001 |
| H-index<sup>b</sup> | 49 (30-89) | <0.001 |  |  |
| Site Specific |  |  |  |  |
| Yes | 77 (31%) | 0.94 |  |  |
| No | 174 (69%) |  |  |  |
| Modality Specific |  |  |  |  |
| Yes | 35 (14%) | 0.45 |  |  |
| No | 216 (86%) |  |  |  |
| Journal Type |  |  |  |  |
| Full OA | 95 (38%) | <0.001 | Reference |  |
| Hybrid OA | 156 (62%) |  | 991 (189) | <0.001 |
| Region |  |  |  |  |
| Europe | 119 (47%) | <0.001 | Reference |  |
| North America | 87 (35%) |  | 838 (186) | <0.001 |
| Other | 45 (%) |  | -307 (213) | 0.15 |

<sup>a</sup>Due to presence of outliers, these variables were log transformed for multivariable linear modelling
<sup>b</sup>Due to collinearity with Impact Factor, the H-index was not included in the multivariable model Abbreviations: APC, article processing charge; IQR, interquartile range; OA, open access; SE; standard error USD, US dollars

Figure 1 describes the distributions of APCs for all identified journals, displayed separately for hybrid OA and full OA journals. Overall, hybrid journals had higher publication costs compared to their full OA counterparts. The median APCs for hybrid and full OA journals were 3260 USD and 1958 USD, respectively. Only 11% of hybrid journals charged less than 2000 USD for OA publication, compared to 59% of full OA journals. Furthermore, 14% of hybrid journals had APCs at least 4000 USD, while only 3% of full OA journals charged this amount.

**Figure 1.**
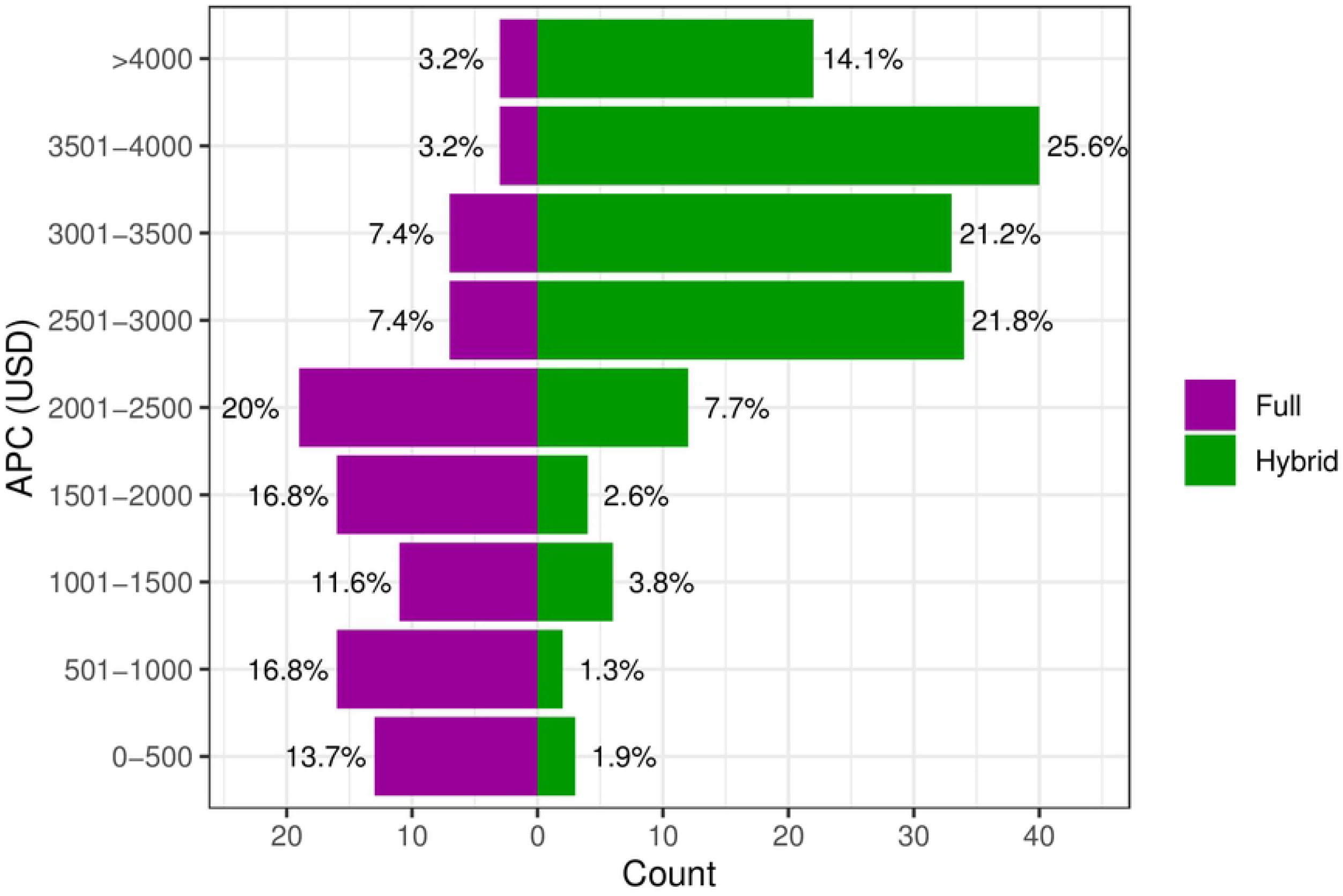
Distribution of article processing charges, displayed separately for journals adopting full and hybrid open access publishing models Abbreviations: APC, article processing charge; USD, US dollars

The associations between APC, journal IF, and region of origin are summarized in Figure 2. Higher journal IF was associated with increased APCs; in general, journals based in Europe and North America were more likely to have higher IF and publication costs. Only 21% and 18% of European and North American journals had IF less than 2, respectively, compared with 42% of journals housed in other regions.

**Figure 2.**
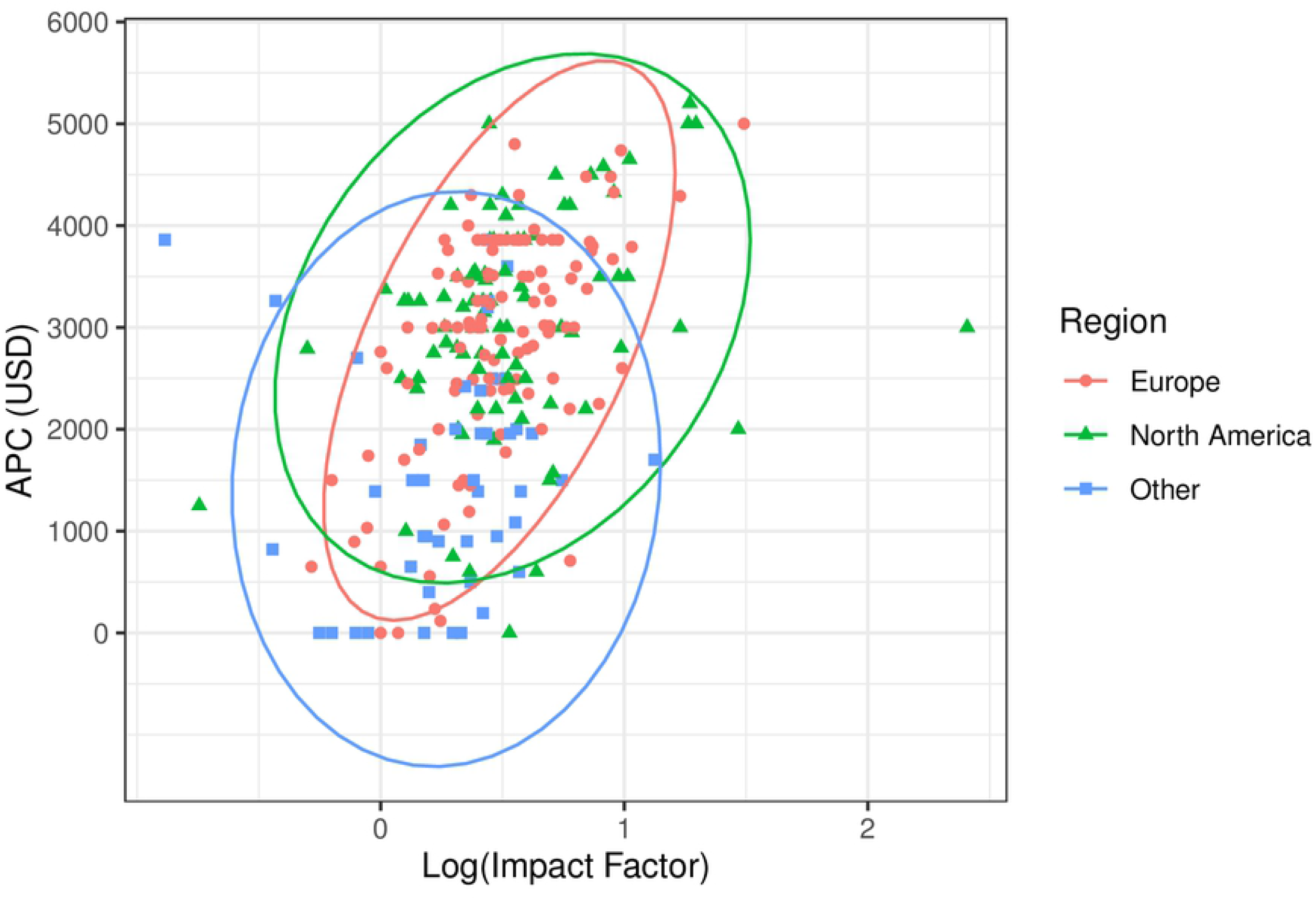
Association between impact factor and article processing charges among journals housed in various global regions Abbreviations: APC, article processing charge; USD, US dollars

## Discussion

In this cross-sectional analysis of OA publication practice among oncology journals, we not only found that a significant portion of journals charged high fees for publication but also identified multiple journal-level factors associated with higher costs. In particular, our data suggest that journals using the hybrid OA model, having higher IF, publishing higher volume of articles, and those housed in North America tend to charge more for authors to publish their article openly. Taken together, these findings suggest that current publication standards in our field may limit sharing of knowledge, particularly among select journals. These data may ideally be used to inform policy-level changes aimed at improving equitable dissemination of knowledge among the oncologic community.

The OA publication model is intended to increase the dissemination of scientific findings by bypassing consumer financial barriers to article access. Indeed, prior research has shown that articles published in an OA forum are more likely to be viewed and downloaded compared to papers limited by a paywall.(4,5) Furthermore, growing evidence has shown that OA articles are more likely to be cited in subsequent peer-reviewed literature, a phenomenon termed the “open access citation advantage,” suggesting increased engagement within the scientific community.(6,7) However, the benefit of open publishing is not limited to the scientific or medical community alone. Particularly within the medical field and oncology more specifically, OA publishing creates an opportunity for improving patient education, advocacy, and shared decision making. Access to peer-reviewed literature may deter patients from seeking information from less reliable sources and may further improve public trust in science and medicine. Finally, because medical research is often supported through public funds, the public may have a strong interest in accessing science in their role as funders, advocates, and research participants.(8)

Plan S is an open access initiative that stipulates research derived from public grant funding must be published in a full and immediate open manner. This enterprise is supported by cOAlition S, a consortium of international research funders and research performing organizations. The objective of Plan S is to not only increase the number of articles published in open access journals, but to ensure public funding directly leads to research that the public can easily access. For full compliance with Plan S, authors must publish in OA journals or platforms, publish in subscription journals and make the articles available OA (but cOAlition S will not financially support OA charges), or publish OA in subscription journals under a transformative arrangement (subscription publishers agree to transition to OA by 2025). In addition, Plan S is working on a Journal Comparison Service, which would increase transparency in open access journals by allowing APCs to be directly compared.

While cOAlition S is already supported by the European Commission and the European Research Council (ERC), the U.S. has not yet adopted these standards and is rapidly becoming an outlier in OA policy. The U.S. continues to rely heavily on traditional publishing methods that require reader subscriptions to access articles, and unlike countries such as Argentina and Canada, have not passed legislation requiring the use of open access publishing methods when publishing federally funded research.

It is worthwhile to note that Plan U (Plan Universal), a research preprint initiative, offers a unique compromise in the realm of open access versus traditional publishing methods. Plan U advocates for all funding agencies to mandate research preprints, which would essentially eliminate the time between submission and official publication. Preprints would be instantaneously uploaded online, which would not only allow for an expedited peer-review process from a larger number of individuals but would give everyone access to new and potentially impactful research.(9) Researchers would still be able to publish in subscription-based journals but would additionally release the manuscripts on preprint websites.

Open publication is still the optimal avenue for the sharing of many, if not all, scientific articles. However, many barriers exist which may limit the ability of researchers and authors to publish their work under current publishing models. Most notably is the presence of publication costs in the form of APCs, which authors must pay themselves in order to publish their work. This can be particularly burdensome for individuals with limited personal or institutional resources, as costs of publication can be quite high. In the present study, we found that the median APC was nearly 3000 USD, with 10% of oncology journals charging more than 4000 USD for OA publication of a single article. Many institutions and academic groups have developed strategies to address APCs, such as pooled funds dedicated specifically for submission fees. Furthermore, some journals have instituted waiver policies for financially vulnerable populations, including submitting authors from low- and middle-income countries. However, the adoption of these policies has not been universal and there is variation in the extent of discount across journals.(10)

Significant publication costs not only create the problem of limiting shareability of the science itself, but they also create a structural framework which predisposes to academic inequity. Authors that are fortunate to have available personal funds or strong institutional support may elect to pursue OA publication and therefore reap the benefits described previously, including improved visibility. In contrast, authors unable to afford OA publication fees may need to publish their work behind a paywall or, worse, may have limited options to share their findings in a scientific journal altogether. Consequently, this may create a cyclical disparity in academic opportunities wherein disadvantaged authors are further limited in their ability to participate in and share scientific research.

The optimal solution to the financial crisis of OA publishing likely will not involve consumer strategies to obtain funds to meet APC requirements but rather focus on reform of publication practice at large. Our study provides initial support that certain journal characteristics may be associated with increased risk of high publication costs; these findings may encourage targeted re-evaluation and reform of APC policies among those select journals.

One of the most important trends identified in this analysis is the association between journal IF and APCs (Figure 2). We found that for every 10-fold increase in IF, there was an estimated increase of 1144 USD in this population (Table). Although IF is an imperfect metric, higher impact journals are often desired forums for publication of scientific work, as they may offer increased article visibility and credibility.(10) Journals with higher IF typically receive higher volume of submissions and may be more selective in editorial acceptance decisions. Such journals therefore also may be afforded the ability to charge higher APCs, as authors may be more willing to pay more to have their article freely accessible in a highly visible and respected environment. Furthermore, it is possible that high impact journals may more often publish work from research groups with greater private or institutional resources and, thus, OA APCs are less limiting among this population of researchers. Regardless of the underlying etiology explaining this trend, the strong association between IF and APCs warrants further exploration with re-evaluation of APC policies particularly among the highest impact oncology journals.

Our analysis also revealed a strong relationship between the type of OA publication model and processing costs (Figure 1). At the time of data collection, the majority (62%) of journals utilized a hybrid model wherein authors are offered the option of publishing OA after paying an APC. We found that hybrid journals tended to have significantly higher OA publication costs, with an estimated difference of 991 USD compared to their full OA counterparts (Table). This trend likely reflects the impact of choice on author submission preferences. For full OA journals publishing exclusively OA papers, authors are not granted any choice and thus every submitting author is required to pay a publication fee. In contrast, with the hybrid model only authors with significant interest in having their article to benefit from open publication and, more importantly, those willing and able to pay the APC will elect for OA publication. As a result, journals adopting the hybrid model may be able to be more stringent in in their APCs without deterring potential submissions. As described above, however, this pattern likely does introduce biases which hinder the ability of authors with limited resources to participate in open science. Therefore, we argue that even though authors are granted choice of participation in the hybrid model, these journals may actually benefit the most from modification of publication standards to be less financially exclusionary of authors from differing backgrounds.

This investigation is limited by several factors related to the study design. First, as with any observational study, evaluation of causal relationships between variables is difficult. However, we did attempt to consider all publicly available data with intuitive potential impact on APCs when generating our multivariable model. Second, some endpoints, such as the proportion of papers published OA in hybrid journals, were of interest for the scope of this investigation but were not readily available online and thus may limit our ability to analyze patterns in author submission preferences. Last, although we did survey all oncology journals listed in the SCImago database, a minority of journals were excluded either due to lack of transparent APC information or non-English language, which may limit the generalizability of our findings.

## Conclusion

OA publication has been shown to improve article visibility compared to traditional subscription-based publication; however, APCs can pose a significant burden to authors and researchers interested in publishing their work in an OA forum. In this cross-sectional observational study of publication practices among oncology journals, we find that APCs were greater in journals with higher IF, more citable documents, those originating in North America, and those utilizing the hybrid OA model. These results warrant further investigation and re-evaluation of publication standards to promote equitable sharing of oncologic research and knowledge.

## Data Availability

Data will be available upon request to the corresponding author.

NA

